# Evoked oscillatory cortical activity during acute pain: Probing brain in pain by transcranial magnetic stimulation combined with electroencephalogram

**DOI:** 10.1101/2024.01.22.24301597

**Authors:** Enrico De Martino, Adenauer Casali, Silvia Casarotto, Gabriel Hassan, Bruno Andry Nascimento Couto, Mario Rosanova, Thomas Graven-Nielsen, Daniel Ciampi de Andrade

## Abstract

Temporal dynamics of local cortical rhythms during acute pain remain largely unknown. The current study used a novel approach based on transcranial magnetic stimulation combined with electroencephalogram (TMS-EEG) to investigate evoked-oscillatory cortical activity during acute pain. Motor (M1) and dorsolateral prefrontal cortex (DLPFC) were probed by TMS, respectively, to record oscillatory power (event-related spectral perturbation and relative spectral power) and phase synchronization (inter-trial coherence) by 63 EEG channels during experimentally induced acute heat pain in 24 healthy participants. TMS-EEG was recorded before, during, and after noxious heat (Acute Pain condition) and non-noxious warm (Control condition), delivered in a randomized sequence. The main frequency bands (α, β1, and β2) of TMS-evoked potentials after M1 and DLPFC stimulation were recorded close to the TMS coil and remotely. Cold and heat pain thresholds were measured before TMS-EEG. Over M1, Acute pain decreased α-band oscillatory power locally and α-band phase synchronization remotely in parietal-occipital clusters compared with non-noxious warm (all P<0.05). The remote (parietal-occipital) decrease in α-band phase synchronization during Acute Pain correlated with the cold (P=0.001) and heat pain thresholds (P=0.023) and to local (M1) α-band oscillatory power decrease (P=0.024). Over DLPFC, Acute Pain only decreased β1-band power locally compared with non-noxious warm (P=0.015). Thus, evoked-oscillatory cortical activity to M1 stimulation is reduced by acute pain in central and parietal-occipital regions and correlated with pain sensitivity, in contrast to DLPFC, which had only local effects. This finding expands the significance of α and β band oscillations and may have relevance for pain therapies.

## INTRODUCTION

Along with structural connections, neuronal assemblies exchange information through oscillatory activity [3]. Neuronal oscillations and their synchronization, concomitantly occurring in different frequency bands, enable information processing across spatially distant brain regions by inter-areal phase-locking. This process creates time windows when information can be integrated concomitantly in sparse neuronal clusters, allowing complexity to emerge [17]. Increasing evidence indicates that individuals with acute and chronic pain exhibit altered electroencephalogram (EEG) [42]. During acute heat pain, the activation of cortical networks resulted in modifications in different frequency bands, including α-band [19], β-band [39], and C-band [52] in healthy individuals. In chronic pain patients, decreased high α-band and low β-band (10-20 Hz) were reported to occur, coupled with an increase in high β-band (20-30 Hz) [36], and related to pain symptoms in peripheral neuropathic pain [54]. While changes in the β-band tend to be confined to frontal cortical regions, changes in the α-band appeared to be more widespread but rather located in central and parietal-occipital cortical regions [19,31,36]. Slowing of the peak of the α rhythm over the sensorimotor cortex has also been suggested as a possible and reliable biomarker of pain sensitivity [20]. However, our understanding of brain oscillations during pain states is mainly based on studies investigating resting-state EEG, not being designed to provide detailed insights into the alterations in oscillatory dynamics of specific cortical regions.

Evoked oscillatory dynamics of a specific brain region can be assessed by recording the EEG responses after perturbating pulses of transcranial magnetic stimulation (TMS-EEG). TMS-EEG measurements can be performed by targeting the primary motor cortex (M1), which is known to have major connections to interoceptive and cognitive networks [3], but also targeting extra-motor areas, such as the dorsolateral prefrontal cortex (DLPFC) [26]. A key feature of TMS-EEG is the ability to transiently affect both the oscillatory power (i.e., event-related spectral perturbation) and phase synchronization (i.e., inter-trial coherence) in the targeted cortical region [4]. While oscillatory power refers to the magnitude of brain oscillations in a specific frequency band and is related to cortical excitability, phase synchronization refers to phase coherence of TMS-evoked potential (TEP) responses over multiple trials [29]. To date, it is unknown how pain influences the evoked-oscillatory cortical activity of cortical areas highly relevant to pain cortical networks, such as M1 [21] and DLPFC [53]. Unveiling these patterns would provide unique and novel models of how specific cortical areas react to pain stimuli locally and how they drive and engage responses in remote areas, which could have major mechanistic and therapeutic implications.

Here, we used TMS-EEG to probe cortical oscillations in M1 and DLPFC during induced acute heat pain and non-painful warm control stimulation in healthy participants. Based on previous studies showing changes in α-band and β-band [19,20,39], we hypothesized that acute heat pain would affect oscillations in the α-band at central regions and remotely in parietal-occipital regions when targeting M1, whereas DLPFC stimulation would affect oscillations in the β-band in the frontal region.

## METHODS

### Participants

In adherence to the Helsinki Declaration, the study was approved by the local ethics committee (Videnskabsetiske Komite for Region Nordjylland: N-20220018) and was registered at ClinicalTrials.gov (NCT05566444). This study is based on original data from a study in which the cortical excitability results have been reported, measured through local peak-to-peak amplitude and slope, and global mean field power [30]. A total of 24 healthy (12 females) right-handed individuals were included (age: 27±5.5 years, weight: 70±14 kg, height: 173±10 cm). All participants were healthy, did not suffer from neuropsychological or other medical conditions, and did not assume any medicaments. Sample size calculations were determined based on previous data exclusively related to local and global cortical excitability results to provide 80% power, type I error rate of 0.05, and type II error rate of 20% [30].

### Experimental protocol

In a single experimental session, eight TMS-EEG blocks were performed by probing two cortical areas relevant for pain modulation (left DLPFC and left M1) under four distinct conditions: Baseline, Acute pain, Non-noxious warm, and Post (Figure 1). The sequence of cortical stimulation areas and the order of Acute pain and Non-noxious warm conditions were randomized among participants to ensure equal distribution and balance between the two groups in terms of participant number. A 5-minute interval separated each block, and a 30-minute break was provided between cortical areas. Pain thresholds for each participant were determined using a thermode (Medoc advanced medical system, Haifa, Israel) with a Peltier-based probe (3 x 3 cm) placed on the right forearm’s volar region. Starting at 32°C, the warm detection threshold (WDT) was determined by the methods of limits. WDT was measured by an increasing temperature (1°C/s) until the participant perceived warm and pressed a stop button. Heat-and cold-pain thresholds (HPT and CPT) were measured similarly by asking the participant to press a stop button when the pain was perceived (average of 3 runs) [44], and approximately 30 seconds separated each measurement. Before experiments, the temperature to be used in the Acute pain and Non-noxious warm stimulation sessions was determined. For Acute pain and Non-noxious warm stimulation, participants indicated the probe temperature needed to produce moderately intense heat pain (Acute pain) and a harmless warm sensation (Non-noxious warm). Beginning at HPT, the probe temperature increased in 1°C increments until participants reported moderately intense heat pain, rated as 5 out of 10 on a numerical scale (0 being no pain, 10 being the worst pain imaginable).

**Figure 1:**
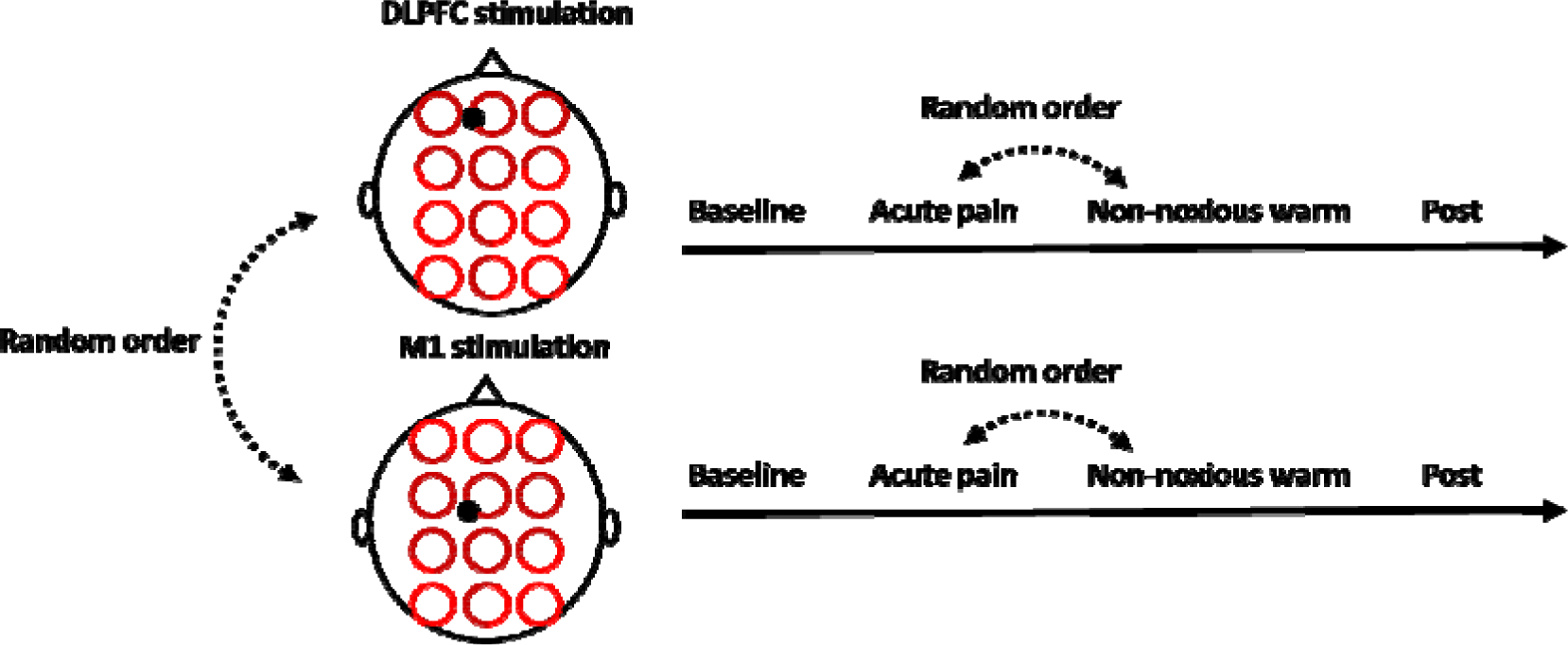
Transcranial magnetic stimulation-electroencephalography was performed in two cortical regions: the dorsolateral prefrontal cortex (DLPFC) and primary motor cortex (M1). Four different conditions were collected for each cortical area: Baseline, Acute pain, Non-noxious warm, and Post.

Temperatures of 45.2±0.7°C were used for Acute pain conditions during TMS-EEG data collection, while 40.2±0.8°C (below HPT) induced the innocuous warm sensation (Non-noxious warm). For Baseline and Post measurements, a 32°C thermode stimulator probe (skin temperature) was used to prevent any thermal sensation. Additional information can be found in De Martino et al. [30].

### Electroencephalographic recordings during transcranial magnetic stimulation

A biphasic stimulator (Magstim Super Rapid^2^ Plus^1^, Magstim Co. Ltd, Dyfed, United Kingdom) and a figure-eight-shaped coil (70 mm, Double Air Film Coil) were used to stimulate DLPFC and M1. TMS-evoked potentials (TEP) were recorded using a TMS-compatible passive electrode cap with 64 electrodes (EASYCAP GmbH, Etterschlag, Germany) placed according to the 10-5 system, with the Cz electrode aligned to the vertex of the head. The electrode impendence was maintained below 5 kΩ during the recordings. Raw signals were amplified and sampled at 4800 Hz (g.HIamp EEG amplifier, g.tec-medical engineering GmbH, Schiedlberg, Austria). The online reference was on a forehead electrode, two electrodes recorded the electrooculogram (EOG) on the lateral side of the eyes, and the ground electrode was situated midway between the eyebrows. To reduce neck and shoulder postural muscle activity, participants sat on an ergonomic chair equipped with dedicated neck support. To minimize oculomotor muscle activity, participants had an easy-to-see fixation spot on the wall. To abolish auditory responses to TMS coil clicks, TMS-click sound masking toolbox (TAAC) with noise-cancelling in-ear headphones (Shure SE215-CL-E Sound Isolating, Shure Incorporated, United States) was used [48]. Finally, to reduce somatosensory sensations from the TMS coil and any EEG electrode movement artefacts, two net caps (GVB-geliMED GmbH, Ginsterweg Bad Segeberg, Germany) with a plastic stretch wrap handle film were applied over the EEG cap.

Using an optical-tracking system, a navigated brain stimulation system (Brainsight TMS Neuronavigation, Rogue Research Inc., Montréal, Canada) calibrated the head of the participant and TMS coil position. The optical-tracking system also generated a 3D brain reconstruction using template MRI (Brainsight software, Rogue Research), scaling to the head of the participant to optimize the reliability of targeting during sessions. The M1 target was identified near the left hemisphere’s hand knob of the central sulcus, where the largest motor-evoked potential (MEP) was recorded by electromyographic electrodes placed on the right first dorsal interosseous muscle (FDI) (i.e., the hand “hot spot”). The resting motor threshold (rMT) was determined as the TMS intensity required to produce MEPs greater than 50 μV in 5 out of 10 trials, with pulses delivered at 0.2 Hz, as measured from the FDI muscle electromyography [46]. Disposable surface silver/silver chloride adhesive electrodes (Ambu Neuroline 720, Ballerup, Denmark) were used to record MEPs in the FDI muscle, placed parallel to the muscle fibers. A reference electrode was positioned on the ulnar styloid process. To avoid sensory-feedback contamination, M1 TMS-evoked potentials were applied below the rMT (90% of rMT) [15]. The DLPFC target was identified in the middle frontal gyrus following Mylius et al. 2013 [37], with the stimulator intensity set at 110% of rMT of the FDI muscle. A real-time visualization tool (rt-TEP) was used to ensure detectable TMS-evoked potentials in both cortical targets [5], allowing minor adjustments of TMS coil orientation across participants to reduce variance and guarantee a minimum of 6 μV in the early peak-to-peak amplitude response in the average of 20 trials in the nearest EEG electrode to DLPFC and M1 targets. The navigated brain stimulation system and rt-TEP were utilized throughout the study to monitor TMS coil location (within 3 mm of cortical targets) and the highest signal-to-noise ratio in EEG recordings. For each condition (Baseline, Acute pain, Non-noxious warm, and Post), approximately 160-180 pulses (∼8 min of TMS stimulation) were administered, with interstimulus intervals randomly jittered between 2600 and 3400 ms to prevent significant reorganization/plasticity processes from interfering with longitudinal TMS/EEG measurements [6].

### Data processing

Data pre-processing was carried out using customized algorithms based on the EEGlab toolbox [10] running on Matlab R2019b (The MathWorks, Inc., Natick, MA, United States). EEG signals were divided into trials of 1600 ms around the TMS pulse (±800 ms with time 0 corresponding to the TMS pulse). The TMS artifact was removed from all EEG recordings by replacing the recording between −2 and 6 ms from the TMS pulse with pre-pulse signal (−11 to −3 ms). Epochs and channels containing noise, eye blinks, eye movements, or muscle artifacts were visually inspected, catalogued, and discarded. Epochs were band-pass filtered (2-80 Hz, Butterworth, 3rd order) and down sampled to 1200 Hz. Channels were re-referenced to the average reference, baseline corrected, and the four conditions (Baseline, Acute pain, Non-noxious warm, and Post) were concatenated. In this merged dataset, independent component analysis (ICA, EEGLAB runica function) was used to eliminate any residual artifacts (i.e., eye blink and TMS artifact). Subsequently, epochs were re-segmented within a ±600 ms time window, and the combined dataset was divided back into the original four conditions. Spherical splines were used to interpolate bad channels [10].

The following TMS-evoked EEG parameters were extracted in the time-frequency domain:

1. The event-related spectral perturbation (ERSP) and relative spectral power (RSP) were calculated to quantify the power amplitude independent of phase. The ERSP allows identifying the changes in power as a function of time and frequency, and RSP provides specific normalized information about the distribution of power across frequency bands.
2. Inter-trial coherence (ITC) was extracted as a measure of phase synchronization.

Time-frequency maps were extracted between 8 and 45 Hz using Morlet wavelets with 3.5 cycles as implemented in the EEGLAB toolbox and previously reported [12,16,45]. ERSP was computed from the time-frequency maps as the ratio of the spectral power of individual EEG trials relative to the pre-stimulus period [13,45]. The significance of ERSP maps with respect to the baseline was assessed by bootstrapping samples from the pre-stimulus period (500 permutations, two-sided comparison, p-value < 0.05 after False Discovery Rate (FDR) correction for multiple comparisons). Mean power spectra were then calculated by averaging significant ERSP values across channels and time-samples. RSP was extracted from the mean power spectra as the percentage of power in a given frequency band [12]. Finally, ITC was calculated by normalizing the complex-valued single-trial time-frequency values by their corresponding moduli and taking the absolute value of the across-trials averaged results. The significance of ITC maps with respect to the baseline was assessed by bootstrapping samples from the pre-stimulus period (500 permutations, one-sided p- value < 0.05 after FDR), and significant ITC values were averaged across channels, time-samples, and frequency bands.

The EEG channels were organized into clusters by averaging individual channels to determine regional neural activity, and ITC, ERSP, and RSP were extracted over a time interval of 6-300 ms after the TMS stimulus for four distinct frequency bands: α (8–12 Hz), β1 (13–24 Hz) and β2 (25- 34 Hz). For the statistical analysis, the absolute change from Baseline was calculated for Acute Pain, Non-noxious warm, and Post. Evoked responses were assessed locally where the TMS pulse was delivered (Figure 2) and remotely in related cortical regions, i.e., those areas typically presenting reverberating oscillations within the natural frequencies of the TMS target. Thus, local responses for M1 probing were assessed in the α-band on the middle centro-frontal cluster (Table 1), while for DLPFC stimulation, local responses were assessed in the β1-band and β2-band on the left/ middle/ right prefrontal clusters (Table 2). Remote effects of M1 probing were also assessed where α-band activity has its natural peak frequency (i.e., posterior-occipital regions [31,50] situated in left/middle/right parieto-occipital clusters – Table 1). Remote prefrontal cluster responses were assessed to control for the expected α-preponderant parietal-occipital cluster responses triggered by M1 probing, while remote responses in parietal-occipital areas were assessed after DLPFC perturbations to control for the expected frontal β-band responses.

**Figure 2:**
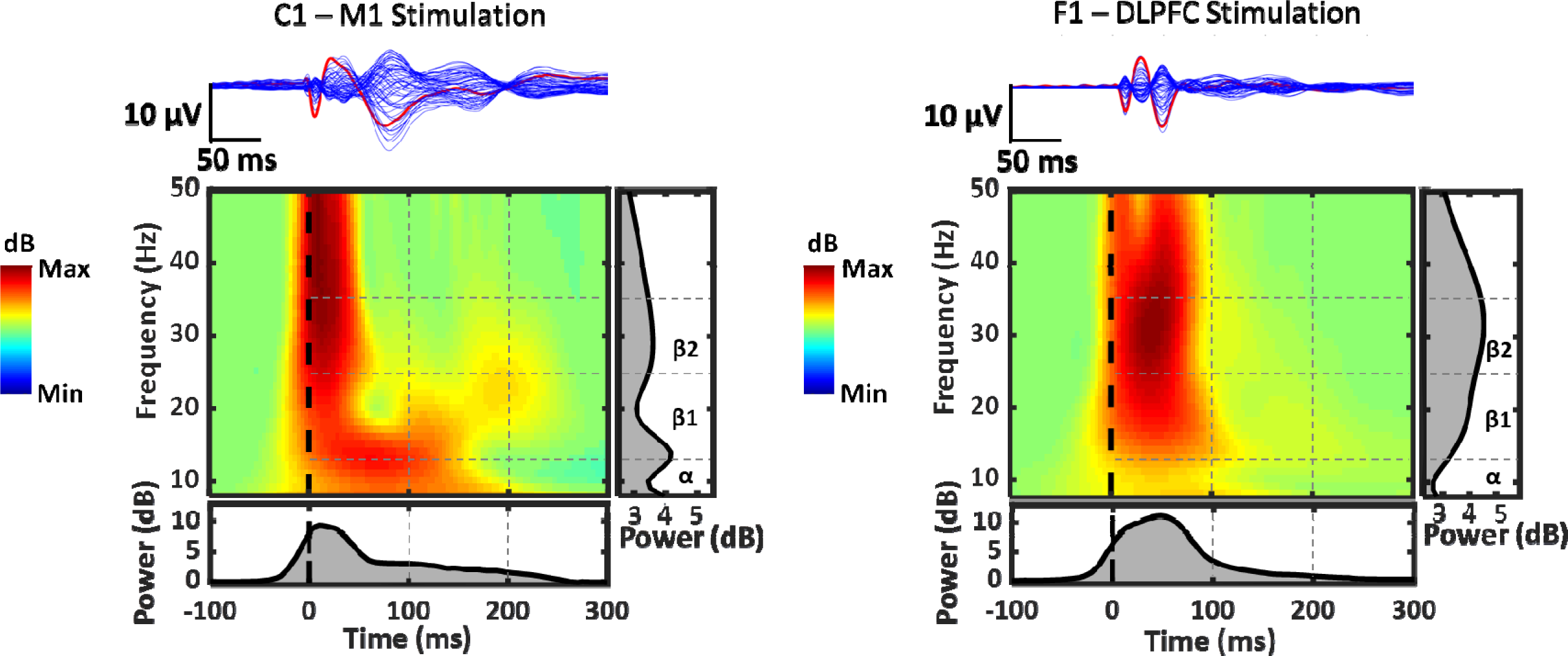
Sample data of transcranial magnetic stimulation (TMS)-evoked potentials recorded with EEG following single pulse stimulation in a representative participant. The figures depict the primary motor cortex (M1) and dorsolateral prefrontal cortex (DLPFC) stimulation with the butterfly plot and topographical maps. The red line corresponds to the C1 and F1 electrodes, and the blue lines correspond to the other 62 channels. The bottom panel shows significant ERSP maps calculated on C1 and F1 electrodes. The gray scale graph plotted at the right depicts the mean power spectrum profile in the α, β1, and β2 bands during the 300 ms after TMS, and the gray scale graph plotted below depicts the mean broadband evoked power during time.

**Table 1:** EEG clusters along with their corresponding electrodes used for M1 stimulation.

| Cluster name | Electrodes |
| --- | --- |
| Middle Centro-Frontal | FCz, Cz, FC1, FC2, C1, C2 |
| Middle Parieto-Occipital | POz, Oz, PO3, PO4 |
| Left Parieto-Occipital | PO7, PO3, O1 |
| Right Parieto-Occipital | PO8, PO4, O2 |

**Table 2:** EEG clusters along with their corresponding electrodes used for DLPFC stimulation.

| Cluster name | Electrodes |
| --- | --- |
| Middle Prefrontal | AFz, Fz, F1, F2 |
| Left Prefrontal | AF7, AF3, F5, F3 |
| Right Prefrontal | AF8, AF4, F6, F4 |

If frequency bands and clusters revealed significant differences between Acute pain and its comparators in the broader 6–300-time interval, then differences in shorter time intervals were explored (i.e., 6-100 ms, 100-200 ms, and 200-300 ms) so that it would be possible to determine whether the significant changes occurred early, middle, or later relative to the probing TMS pulse.

Importantly, reduced α power during acute experimental pain has been reported in resting state-EEG experiments [7,8]. Therefore, we planned a supplementary assessment to rule out a potential confounding pain-related decrease in α power before the delivery of the probing TMS pulse (time interval −600 ms to −10 ms). The α power was calculated by applying the Fast-Fourier Transform to the spontaneous EEG of the pre-TMS stimulus for each individual trial and then averaging the resulting power spectrum across trials. This analysis was conducted exclusively in the electrode clusters exhibiting statistical differences between Acute pain and other conditions.

### Statistical analysis

Statistical analysis was performed with the Statistical Package for Social Sciences (SPSS, version 25; IBM, Chicago, United States). Results were presented as means and standard deviation with a two-sided 5% significance level set for statistical significance and if not otherwise stated. All data from thermal stimulation (Acute pain and Non-noxious warm) and for Post measures are reported as absolute changes from the Baseline. All measurements were assessed by visually examining histograms and Shapiro–Wilk tests. Due to several non-normally distributed parameters, Friedman tests were used to analyze ESRP, RSP, and ITC for each band frequency and cluster, as well as for the α power before TMS stimulation. Post-hoc analyses were performed with Wilcoxon’s multiple comparison tests, and Bonferroni correction was applied when necessary. To determine whether functional connectivity changes during Acute pain were associated with HPT and CPT, Spearman’s rank correlation analyses were conducted between HTP and CPT and the absolute changes from the Baseline of the ERSP, RSP, and ITC during Acute pain. Only significant changes in specific frequency bands and clusters were considered for correlations. Finally, Spearman’s rank correlation analysis was performed on the absolute changes from the Baseline between α ERSP and α ITC from M1 stimulation to investigate whether local α changes correlated with remote α changes since both were significantly changed during Acute pain.

## RESULTS

All volunteers participated in all experimental sessions and underwent all assessments. No adverse events related to TMS-EEG or thermal stimulations were present.

### Spectral power changes

Upon M1 probing, a significant decrease in the power of α-band ERSP was found locally in the middle centro-frontal cluster (Chi-square = 10.083; P = 0.006) as well as in the α-band RSP (Chi-square = 10.750; P = 0.005) at the time interval 6-300 ms (non-normalized parameters are reported in Supplementary Table 1). Post-hoc analysis revealed a significant decrease during Acute Pain compared to Non-noxious warm in α-band ESRP (P = 0.003; Bonferroni-corrected) (Figure 3) and in α-band RSP (P = 0.009; Bonferroni-corrected) (Figure 4A). Shorter time intervals were then explored for changes in α-band ERSP after M1 probing in the middle centro-frontal cluster. Significant differences were found at the time interval 6-100 ms (Chi-square = 6.750; P = 0.034), 100-200 ms (Chi-square = 11.083; P = 0.004), and 200-300 ms (Chi-square = 10.333; P = 0.006) so that for all three-time intervals post-hoc analysis revealed a significant decrease in Acute pain compared to Non-noxious warm (6-100 ms, P = 0.003; 100-200 ms, P = 0.006, 200-300, P = 0.012; all Bonferroni-corrected) (Supplementary Figure 1). A significant difference was also detected in later latencies for α-band RSP (200-300 ms: Chi-Square = 7.583; P = 0.023), but post-hoc analysis did not detect any difference between conditions (P > 0.05; Bonferroni-corrected).

**Figure 3:**
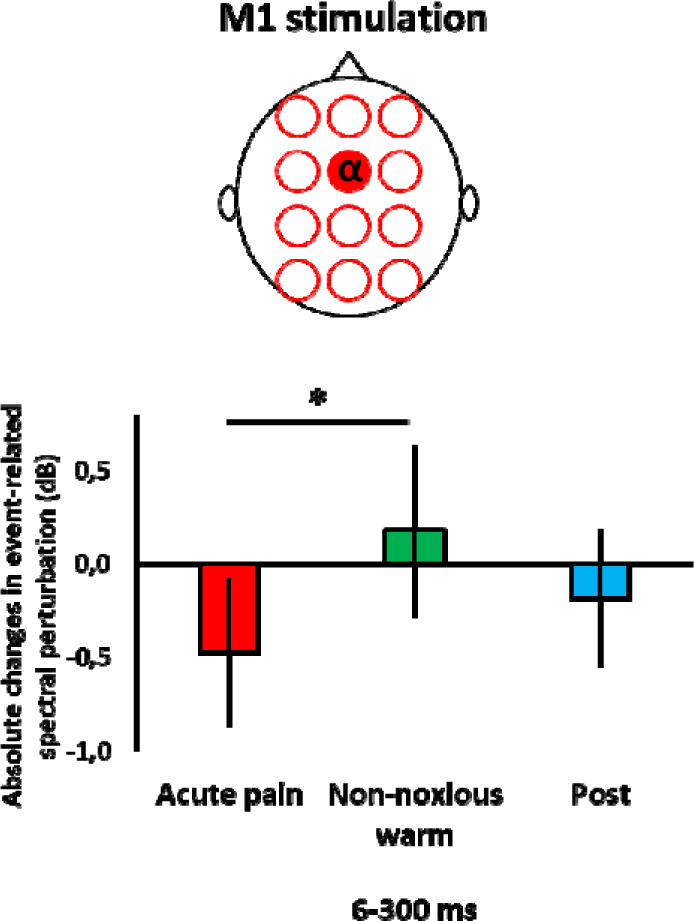
The event-related spectral perturbation absolute changes from Baseline (mean and 95% confidence interval) in the middle centro-frontal cluster are shown during Acute Pain, Non-noxious warm, and Post (Wilcoxon test * P <0.05 – Bonferroni corrected).

**Figure 4:**
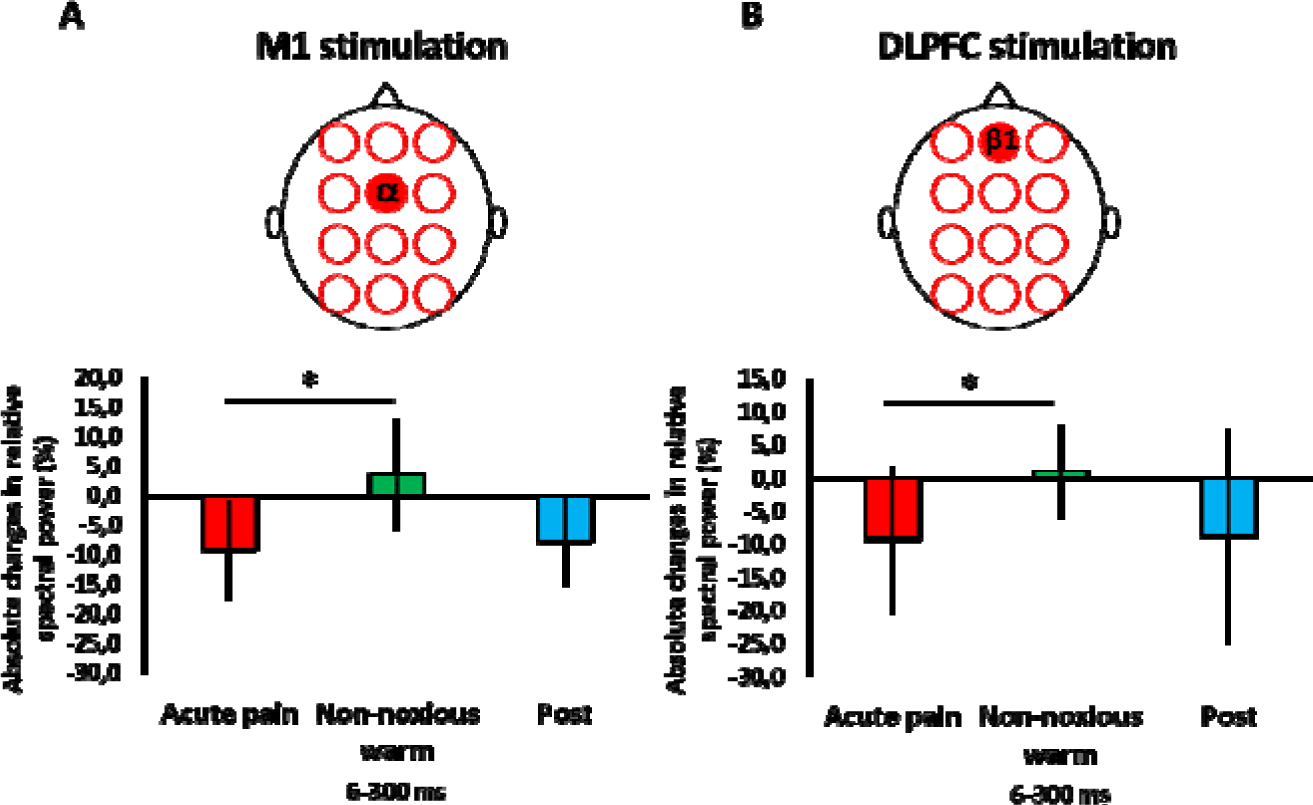
The relative spectral power absolute changes from Baseline (mean and 95% confidence interval) in the middle prefrontal cluster are shown during Acute Pain, Non-noxious warm, and Post (Wilcoxon test * P <0.05 – Bonferroni corrected).

Upon DLPFC probing, a reduction in power in the β1-band RSP was found locally (middle prefrontal cluster) (Chi-square = 12.250; P = 0.002) at the time interval 6-300 ms. Post-hoc analysis revealed a decrease in Acute pain compared to Non-noxious warm (P = 0.015; Bonferroni-corrected) (Figure 4B). When shorter time intervals were analyzed for the β1-band RSP, a difference was found to be only localized at the time interval 6-100 ms (Chi-Square: 9.250; P = 0.010). Post-hoc analysis revealed a reduction in RSP in the β1-band power during Acute pain compared to Non-noxious warm (P = 0.018; Bonferroni-corrected) (Supplementary Figure 2). No local differences were detected in the ERSP (all P > 0.05). Probing of the DLPFC did not lead to significant ERSP and RSP changes in the α-band, β1-band, and β2-band on the left and right prefrontal clusters (all P > 0.05 - non-normalized parameters are reported in Supplementary Table 2).

### Absence of confounding pre-probing alpha power changes

The pre-planned sensitivity analyses confirmed that the decrease in α-band ERSP described above was not significantly present before the TMS in the middle centro-frontal cluster. The spontaneous pre-TMS pulse α-band power was 0.88±0.62 dB for Baseline, 0.94±0.62 dB for Acute pain, 0.94±0.63 dB for Non-noxious warm, and 0.90±0.60 dB for Post. These differences were not statistically different (Chi-square = 0.583; P = 0.747).

### Phase synchronization changes

Upon M1 probing, a significant reduction was found in ITC in the α-band locally at the time interval of 6-300 ms. Remote reductions in ITC were significant in parieto-occipital regions: the right (Chi-square = 12.250; P = 0.002), middle (Chi-square = 13.583; P = 0.001), and left parieto-occipital (Chi-square = 6.750; P = 0.034) clusters. Post-hoc analysis confirmed a reduction in α- band ITC after Acute pain compared to Non-noxious warm in all three EEG clusters (right parieto-occipital cluster, P = 0.006; middle parieto-occipital cluster, P = 0.003, and left parieto-occipital cluster, P = 0.021; all Bonferroni-corrected) and between Acute pain and Post condition within the right (P = 0.009; Bonferroni-corrected) and middle (P = 0.012; Bonferroni-corrected) parieto-occipital clusters (Figure 5). The reductions in α-band ITC occurred in both short and middle latencies intervals in the right parieto-occipital channel clusters. For the right parieto-occipital cluster, we found a reduction in the following time intervals: 6-100 ms (Chi-square = 11.516; P = 0.003) and 100-200 ms (Chi-square = 9.979; P = 0.007). In both two-time intervals, post-hoc analysis revealed a decrease in Acute pain compared to Non-noxious warm (6-100 ms, P = 0.009; 100-200 ms, P = 0.006; all Bonferroni-corrected) and in Acute pain compared to Post condition (6- 100 ms, P = 0.006; 100-200 ms, P = 0.012; all Bonferroni-corrected) (Supplementary Figure 3). Within the left and middle parietal-occipital clusters, the α-band ITC showed significant decreases in Acute pain compared to Non-noxious warm in the later time interval (200-300ms). No change was found in the local middle centro-frontal clusters (Chi-square = 0.750; P = 0.687) (non-normalized parameters are reported in Supplementary Table 3).

**Figure 5:**
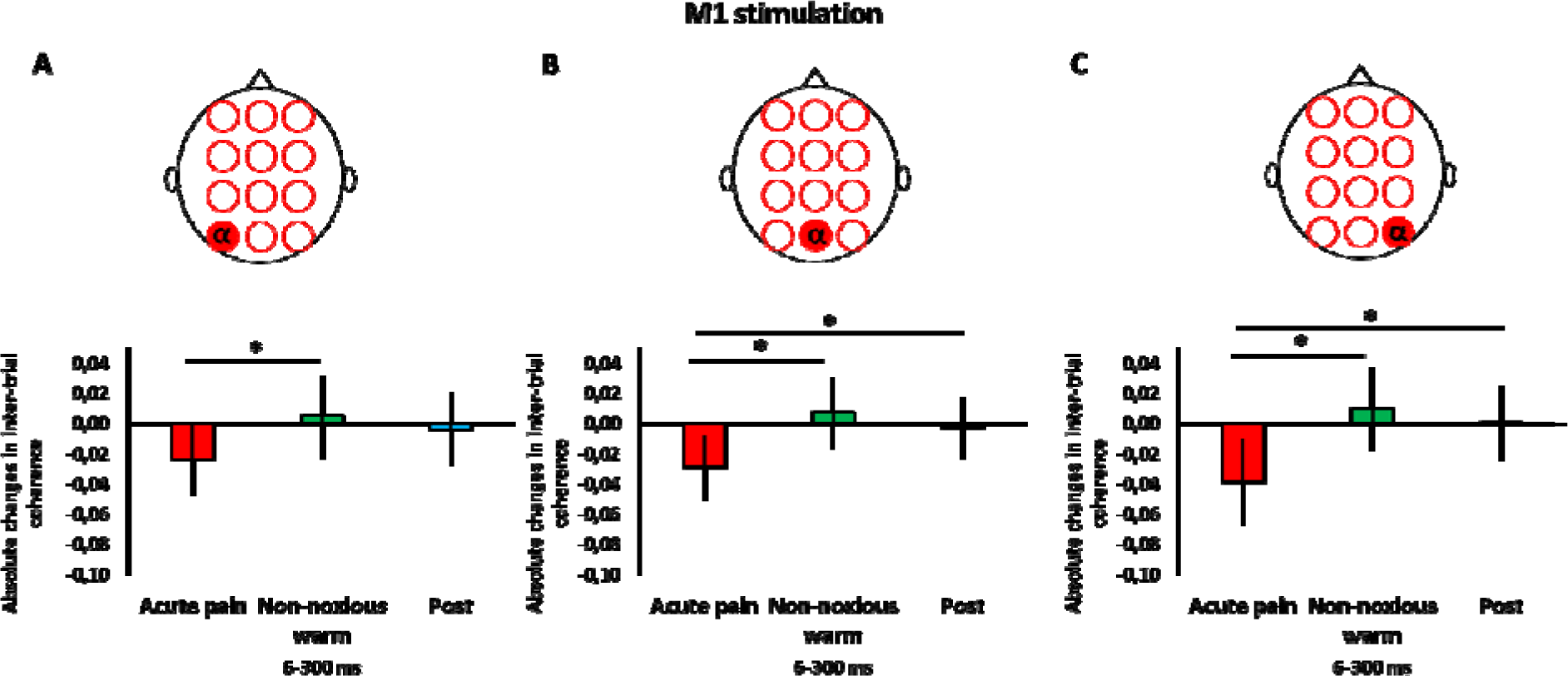
The inter-trial coherence absolute changes from Baseline (mean and 95% confidence interval) in the left (A), middle (B), and right (C) parieto-occipital clusters are shown during Acute Pain, Non-noxious warm, and Post (Wilcoxon test * P <0.05 – Bonferroni corrected).

Probing of the DLPFC did not lead to significant ITC changes in the α-band, β1-band, and β2-band on the left, middle, and right prefrontal clusters (all P > 0.05 - non-normalized parameters are reported in Supplementary Table 4).

### Correlations

During Acute pain, reduction in α-band ITC significantly correlated with cold (rho = 0.638, P = 0.001 - Figure 6A) and heat pain thresholds (rho = −0.463, P = 0.023 - Figure 6B) in earlier latencies (6-100 ms) after M1 stimulation. Upon M1 probing, reduction in α-band ITC locally under Acute pain significantly correlated with the decreases in α-band ERSP in earlier latencies (rho = 0.459, P = 0.024) (Figure 6C). Upon DLPFC probing, no correlations were found between thermal thresholds or local reductions in β1-band power during Acute pain.

**Figure 6:**
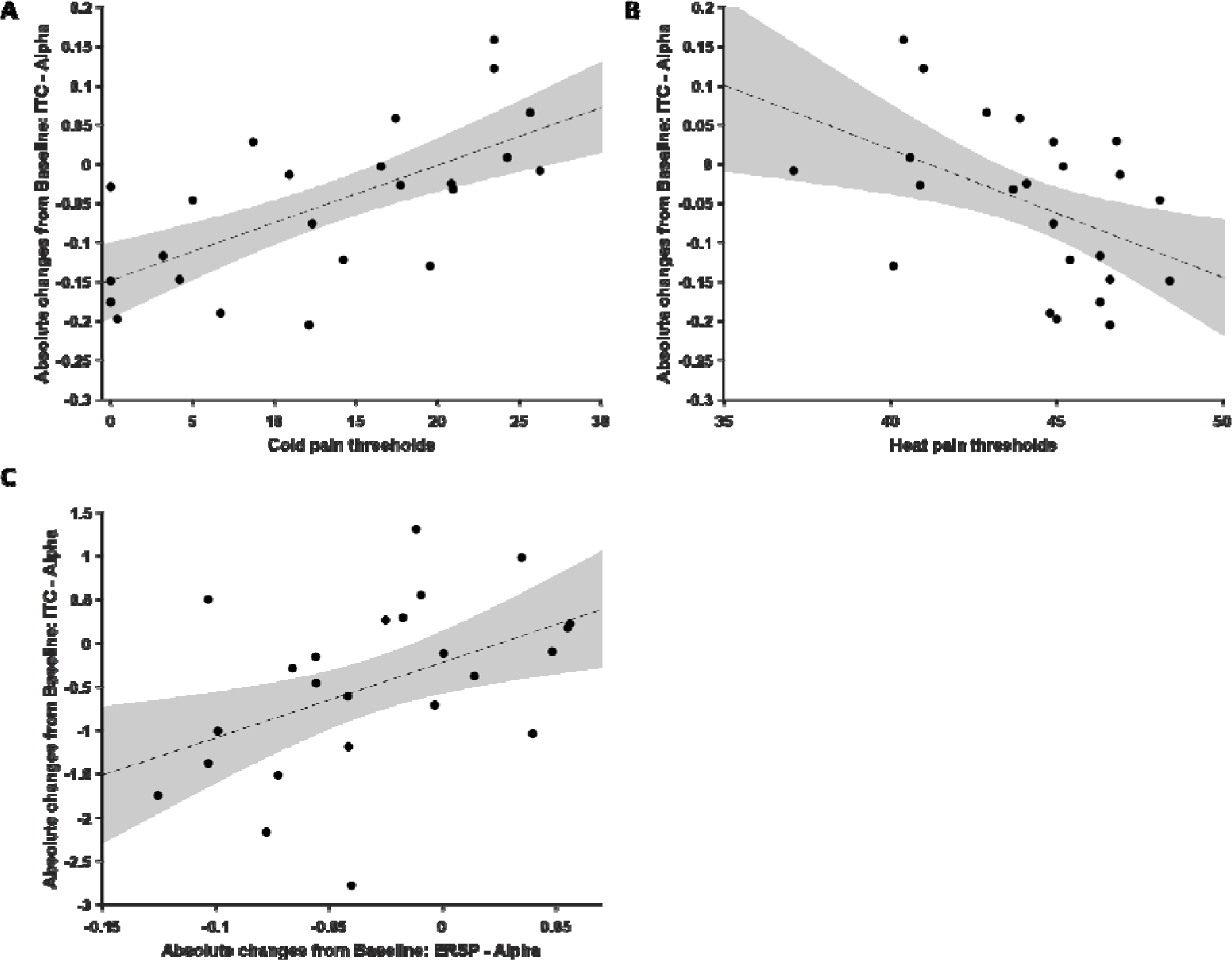
Correlation of the α inter-trial coherence (ITC) responses within the right parieto-occipital cluster from M1 stimulation during Acute Pain, expressed as absolute changes from Baseline. This correlation was measured against Cold (**A**) and Heat Pain Thresholds (**B**) in the time interval of 6-100 ms. **C)** Correlation of the α ITC responses within the middle parieto-occipital cluster from M1 stimulation during Acute Pain, expressed as absolute changes from Baseline. This correlation was measured against α event-related spectral perturbation (ERSP) within the middle centro-frontal cluster in the time interval of 6-100 ms.

## DISCUSSION

The current study investigated the effects of acute pain on evoked oscillatory cortical activity by applying single-pulse TMS to two distinct cortical regions. In terms of power dynamics, acute pain resulted in power reductions localized at clusters near the respective stimulation targets and within specific frequency bands: we observed local decreases within the α-band under M1 stimulation and within the β1-band during DLPFC probing. In terms of phase dynamics, acute pain led to a decrease in α-band synchronization in parietal-occipital clusters under M1 stimulation. Notably, these changes in α-band ITC correlated with thermal pain thresholds, suggesting a potential interaction between trait pain perception and dynamic acute pain-related disengagement of posterior areas, probably via corticothalamic loops.

Traditional resting-state EEG has demonstrated power amplitude and peak frequency changes across various bands in chronic pain patients [36] and healthy participants during experimental pain [38]. In the presence of neuropathic pain, enhanced 0 and high-{] power were described, as well as a decrease in the high a and low-{] power [50]. Furthermore, a shift toward lower peak α frequency was also described in patients affected by neuropathic pain patients [56]. Studies using tonic painful heat stimuli reported a decrease in peak α frequency oscillations in parietal regions [40], a decrease in α and β oscillations in the central region [38,41,57], while faster frequency oscillations power in the middle prefrontal cortex were seen to increase [52]. While these results have offered valuable insights into pain mechanisms, resting-state EEG has limited capability in probing the excitability and connectivity of specific cortical circuits. In the current study, a different methodological approach has been applied, taking advantage of TMS-evoked EEG oscillations, which reflect both local and remote activations from connected populations with various electrophysiological properties as well as the reactivity of the neuronal population at the stimulation site [32,45]. The key finding of the present study was that the alteration in power dynamics during acute pain depends on the cortical region engaged by the TMS stimulation. M1 and DLPFC are part of two different structural and functional connectivity arrangements and are hubs in different brain networks [34]. Dissimilarities between M1 and DLPFC have been described in the spatial-temporal dynamics of M1 activity propagations after TMS stimulation to M1: after the engagement of the stimulation target, activity spreads to more parietal locations via corticocortical volleys from M1 to S1, and to the opposite hemisphere, via the corpus callosum [27]. Differently, the left DLPFC TMS has been described to activate the local stimulation area, as well as the opposite prefrontal cortex [28]. Accordingly, the present results showed that acute pain entrained frequency-specific changes in power depending on the network being probed. The fact that acute pain caused a reduction in power in the α-band after M1 and β-band after DLPFC stimulations can be interpreted according to the natural frequency framework [45]. This means that the main oscillatory frequency evoked by a probing pulse of TMS would be the dominant frequency naturally occurring on that specific cortical area at rest, being the α-band for sensorimotor (Mu rhythm) and β-band for premotor ones. The reduction in power on both targets during acute pain would be in line with previous data showing decreased corticospinal excitability during acute pain [2]. It is possible that local increases in thresholds (i.e., lower excitability) would allow these regions to disengage from their current motor/cognitive processing, thus allowing for plasticity-driven reorganizational changes necessary to respond to acute pain. This aligns with corticospinal excitability modifications in patients with chronic pain, where motor thresholds are rarely abnormal, and plastic changes are more frequently related to intracortical GABA and glutamate-dependent changes [35].

Another main finding of the present study was the reduction of posterior α-band synchronization during acute pain when M1 was probed. While α-band ESRP was locally reduced at the stimulation site in the motor region, decreased α-band ITC did not occur locally. Instead, it took place over remote parietal-occipital electrodes. These changes were more pronounced at early latencies (<100ms) after the M1 pulse and contralaterally. It has been shown that the thalamus acts as the primary pacemaker for α oscillations, with the pulvinar [49] and lateral geniculate nucleus [24] preferentially driving the α rhythm. However, other studies indicate that α waves propagate from higher-to lower-order areas in both the sensorimotor and posterior cortices (from the associative cortex towards the primary cortex) and then to the thalamus, likely via short-range supragranular feedback projections [23]. Intracranial recordings have shown that cortical pyramidal cells modulate excitability and create synchronized feedback loops to the thalamus, leading to highly coherent oscillations [22] and supporting the idea that sensorimotor and posterior cortices play a role in initiating and coordinating oscillations generated within the thalamus [11]. This process involves cortex-thalamus-cortex loops, potentially explaining the generation of large-scale coherent oscillations within the thalamocortical system [18]. We found that M1 stimulation during acute pain decreases the expected parieto-occipital phase synchronization of ongoing rhythmic activity. The intensity of this effect correlated with both the heat and cold pain thresholds of participants, which is one of the few correlations between connectivity metrics and individual trait nociceptive thresholds reported to date. These findings suggest that during acute pain, M1 engages less intensely distant phase synchronization (i.e., lower ITC) in those healthy participants with “trait” higher pain thresholds (i.e., broader non-noxious temperature limen between cold and heat pain thresholds). Therefore, individuals with lower pain sensitivity traits (i.e., higher thermal pain thresholds) exhibit lower inhibitory effects of acute pain in α-band ITC. These correlations reached moderate strength for shorter latencies. Analogously, in the visual system, the accurate perception of the temporal sequence of visual events depends on the phases of the α rhythm [33]. It was also known that sensorimotor networks oscillate at 10-20 Hz [25], which are the frequencies shown to reduce pain intensity in repetitive TMS trials targeting M1 [22], and which is according to Hebbian models [51].

There are limitations that should be considered in interpreting these findings. Firstly, this study did not evaluate the saliency of the non-noxious warm stimulus, which could influence the results. The non-noxious warm condition was utilized as a control to provide comparable sensory inputs to the forearm without inducing pain. To mitigate this limitation, we delivered non-noxious warm stimuli on similar perceptual intensity as acute heat pain [55]. However, it is important to acknowledge that acute heat pain and warm non-noxious stimuli may engage saliency systems differently. While it has been argued that heightened salience is an intrinsic component of pain, a control situation with matched salience intensity delivering a different sensory stimulus could be potentially useful. Secondly, although the current study focused on two major cortical targets, namely M1 and DLPFC, pain engages numerous other cortical regions not probed here. Future research should explore targets like the parietal cortex or deep cortical areas like the posterior insula cortex, which are essential in pain processing [14]. Expanding the cortical targets examined using TMS-EEG can give a more comprehensive understanding of pain mechanisms. Thirdly, several similar outcomes have been analyzed since this study was the first to delve into their exploration in the context of pain. However, due to the limited sample size, only prominent effects could be detected, and more subtle effects are likely to have been overlooked. Furthermore, utilizing multiple corrections might inadvertently neglect potentially significant findings and incur type-II errors [47]. Fourthly, source modeling analysis was not applied in the current study, which could have enabled a more fine-grained localization of cortical areas involved in pain-related neural activity. Finally, cortical responses induced by TMS can be affected by auditory and somatosensory responses [1]. To mitigate this, a control condition with the non-noxious warm stimulus was included and compared differences based on changes from baseline and post-stimulation phases. Additionally, we analyzed short-time intervals as auditory and somatosensory responses predominantly influence the 100-200 ms range [43] despite implementing measures to minimize their impact [5,48].

In conclusion, our results indicated that TMS stimulations to M1 during acute pain drive frequency-specific remote phase synchronization effects, which correlated with nociceptive thresholds and were qualitatively and quantitatively different from the responses seen after DLPFC probing. Our findings likely expand the significance of α-band and {]-band oscillations in perceptual processes to now include nociception.

## Data Availability

All data produced in the present study are available upon reasonable request to the authors

