## Supplementary tables and figures for "Evoked oscillatory cortical activity during acute pain: Probing brain in pain by transcranial magnetic stimulation combined with electroencephalogram"

**SUPPLEMENTARY MATERIAL**

**Supplementary figure 1**

The ERSP absolute changes from Baseline (mean and 95% confidence interval) are shown during Acute Pain, Non-noxious warm, and Post (Wilcoxon test * P <0.05) at the time intervals 6-100 ms, 100-200 ms, and 200-300 ms.

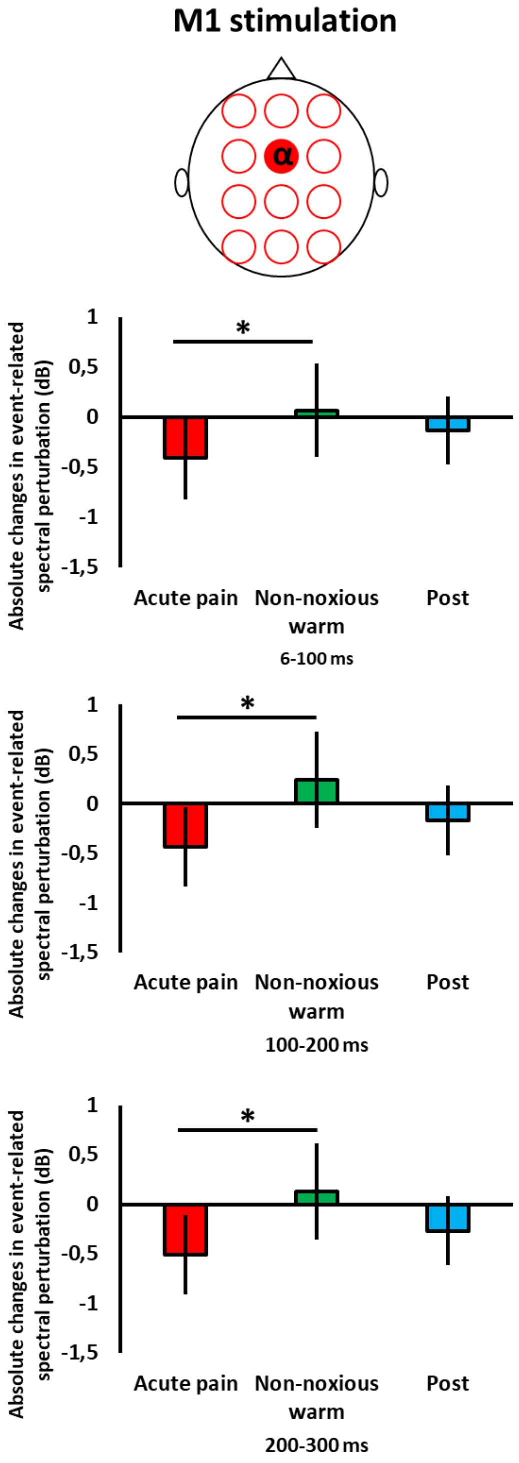

**Supplementary figure 2**

The RSP absolute changes from Baseline (mean and 95% confidence interval) are shown during Acute Pain, Non-noxious warm, and Post (Wilcoxon test * P <0.05) at the time interval 6-100 ms.

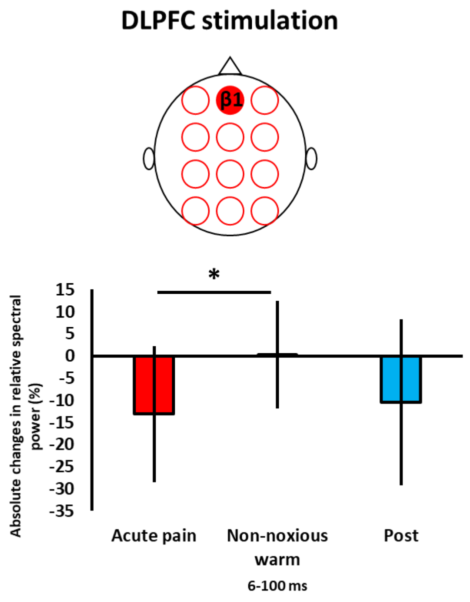

**Supplementary figure 3**

The ITC absolute changes from Baseline (mean and 95% confidence interval) are shown during Acute Pain, Non-noxious warm, and Post (Wilcoxon test * P <0.05) at the time intervals 6-100 ms, 100-200 ms, and 200-300 ms. RPO - Right Parieto-Occipital; MPO - middle Parieto-Occipital; LPO - left Parieto-Occipital

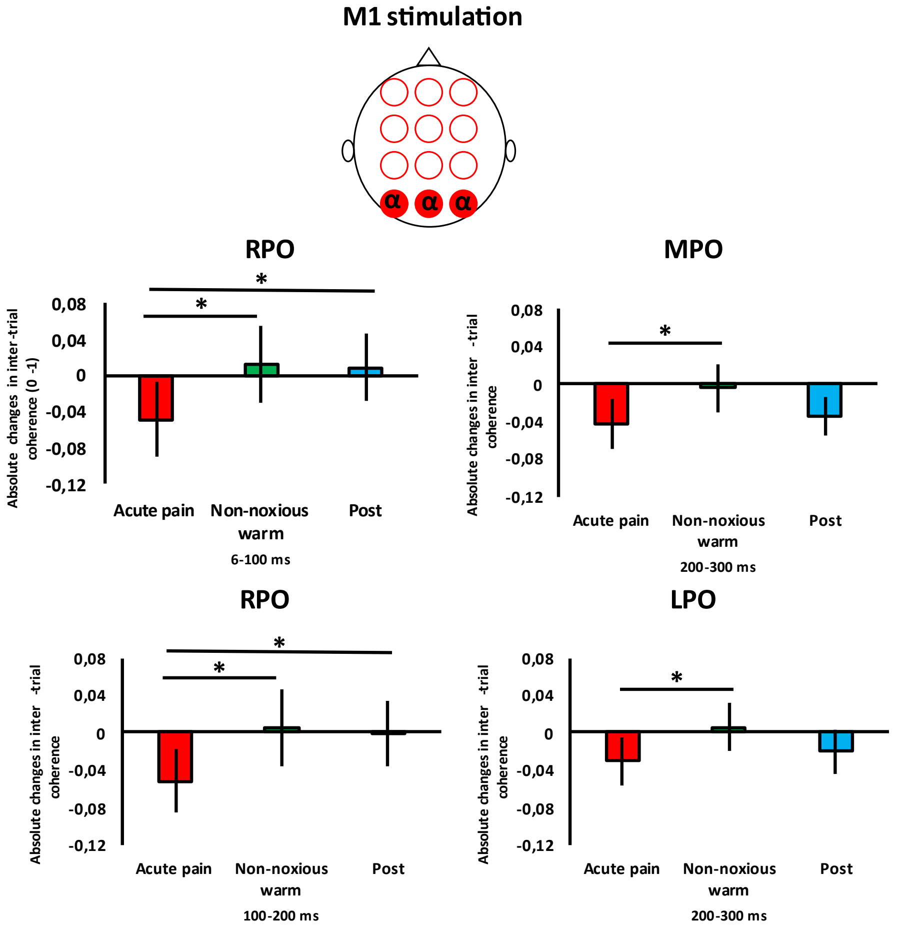

**Supplementary table 1**

Mean (± standard deviation) event-related spectral perturbation (ERSP) during primary motor cortex (M1) stimulation in EEG clusters during 6-300 ms.

| Cluster | Band | ERSP during M1 stimulation – Time interval: 6-300 ms | | | |
|---|---|---|---|---|---|
| | | Baseline | Acute pain | Non-noxious warm | Post |
| Middle Centro-Frontal | α | 1.53±1.82 | 1.06±1.43 | 1.71±1.81 | 1.35±1.73 |
| | β1 | 1.50±0.77 | 1.36±0.91 | 1.36±0.92 | 1.49±1.02 |
| | β2 | 1.62±0.96 | 1.61±1.07 | 1.57±1.09 | 1.81±1.18 |
| left Parieto-Occipital | α | 0.55±0.84 | 0.39±0.57 | 0.50±0.82 | 0.34±0.74 |
| | β1 | 0.57±0.46 | 0.45±0.38 | 0.45±0.44 | 0.50±0.43 |
| | β2 | 0.55±0.37 | 0.52±0.38 | 0.51±0.49 | 0.63±0.49 |
| Middle Parieto-Occipital | α | 0.47±0.77 | 0.37±0.52 | 0.46±0.75 | 0.29±0.55 |
| | β1 | 0.52±0.42 | 0.37±0.28 | 0.39±0.34 | 0.39±0.38 |
| | β2 | 0.43±0.31 | 0.45±0.29 | 0.44±0.37 | 0.51±0.42 |
| Right Parieto-Occipital | α | 0.38±0.69 | 0.26±0.61 | 0.37±0.81 | 0.16±0.49 |
| | β1 | 0.39±0.45 | 0.25±0.27 | 0.26±0.33 | 0.28±0.40 |
| | β2 | 0.29±0.33 | 0.33±0.29 | 0.29±0.33 | 0.37±0.42 |

**Supplementary table 2**

Mean (± standard deviation) event-related spectral perturbation (ERSP) during dorsolateral prefrontal cortex (DLPFC) stimulation in EEG clusters during 6-300 ms.

| Cluster | Band | ERSP during DLPFC stimulation – Time interval: 6-300 ms | | | |
|---|---|---|---|---|---|
| | | Baseline | Acute pain | Non-noxious warm | Post |
| Middle Prefrontal | α | 0.86±1.47 | 0.65±1.16 | 0.76±1.34 | 0.73±1.22 |
| | β1 | 1.84±1.74 | 1.51±1.64 | 1.85±1.79 | 1.73±1.63 |
| | β2 | 2.25±1.96 | 2.12±2.06 | 2.29±2.12 | 2.43±2.32 |
| left Prefrontal | α | 0.67±1.19 | 0.47±0.99 | 0.61±1.18 | 0.62±1.16 |
| | β1 | 1.39±1.56 | 1.17±1.51 | 1.39±1.65 | 1.39±1.67 |
| | β2 | 1.81±1.90 | 1.73±2.10 | 1.83±2.06 | 1.99±2.31 |
| Right Prefrontal | α | 0.69±1.30 | 0.56±1.10 | 0.59±1.16 | 0.61±1.03 |
| | β1 | 0.99±1.02 | 0.82±0.87 | 0.94±0.96 | 0.91±0.82 |
| | β2 | 0.95±0.97 | 0.99±0.92 | 0.93±1.02 | 1.08±1.18 |

**Supplementary table 3**

Mean (± standard deviation) inter-trial coherence (ITC) during primary motor cortex (M1) stimulation in all clusters during 6-300 ms.

| Cluster | Band | ITC during M1 stimulation – Time interval: 6-300 ms | | | |
|---|---|---|---|---|---|
| | | Baseline | Acute pain | Non-noxious warm | Post |
| Middle Centro-Frontal | α | 0.35±0.16 | 0.31±0.17 | 0.34±0.17 | 0.34±0.17 |
| | β1 | 0.24±0.08 | 0.24±0.08 | 0.24±0.09 | 0.25±0.09 |
| | β2 | 0.22±0.07 | 0.23±0.08 | 0.21±0.07 | 0.22±0.07 |
| left Parieto-Occipital | α | 0.19±0.13 | 0.16±0.12 | 0.20±0.13 | 0.19±0.13 |
| | β1 | 0.16±0.07 | 0.16±0.06 | 0.16±0.06 | 0.16±0.07 |
| | β2 | 0.14±0.05 | 0.14±0.06 | 0.14±0.06 | 0.14±0.07 |
| Middle Parieto-Occipital | α | 0.18±0.11 | 0.15±0.10 | 0.19±0.12 | 0.18±0.11 |
| | β1 | 0.14±0.05 | 0.13±0.06 | 0.14±0.06 | 0.14±0.05 |
| | β2 | 0.12±0.04 | 0.13±0.05 | 0.12±0.04 | 0.13±0.05 |
| Right Parieto-Occipital | α | 0.19±0.11 | 0.15±0.10 | 0.20±0.13 | 0.19±0.10 |
| | β1 | 0.12±0.05 | 0.11±0.06 | 0.11±0.07 | 0.11±0.06 |
| | β2 | 0.10±0.04 | 0.10±0.05 | 0.11±0.04 | 0.11±0.05 |

**Supplementary table 4**

Mean (± standard deviation) inter-trial coherence (ITC) during dorsolateral prefrontal cortex (DLPFC) stimulation in EEG clusters during 6-300 ms.

| Cluster | Band | ITC during DLPFC stimulation – Time interval: 6-300 ms | | | |
|---|---|---|---|---|---|
| | | Baseline | Acute pain | Non-noxious warm | Post |
| Middle Prefrontal | α | 0.25±0.14 | 0.21±0.12 | 0.22±0.11 | 0.21±0.12 |
| | β1 | 0.24±0.10 | 0.22±0.10 | 0.22±0.09 | 0.22±0.08 |
| | β2 | 0.22±0.07 | 0.21±0.07 | 0.21±0.06 | 0.21±0.08 |
| left Prefrontal | α | 0.21±0.13 | 0.19±0.11 | 0.20±0.10 | 0.18±0.10 |
| | β1 | 0.18±0.08 | 0.16±0.07 | 0.18±0.07 | 0.17±0.08 |
| | β2 | 0.18±0.05 | 0.17±0.05 | 0.18±0.05 | 0.18±0.06 |
| Right Prefrontal | α | 0.27±0.19 | 0.26±0.17 | 0.27±0.17 | 0.26±0.17 |
| | β1 | 0.18±0.10 | 0.16±0.08 | 0.16±0.08 | 0.16±0.08 |
| | β2 | 0.15±0.06 | 0.15±0.06 | 0.15±0.06 | 0.14±0.06 |
